# The impact of intracranial pressure telesensors: an observational propensity matched-control analysis of service demand and costs

**DOI:** 10.1101/2023.08.01.23293079

**Authors:** Anand S Pandit, Muhammad A Kamal, Gerda Reischer, Yousif Aldabbagh, Mohammad Alradhawi, Faith MY Lee, Priya P Sekhon, Eleanor M Moncur, Ptolemy DW Banks, Simon Thompson, Lewis Thorne, Laurence D Watkins, Ahmed K Toma

**Affiliations:** Victor Horsley Department of Neurosurgery, National Hospital for Neurology & Neurosurgery, London, UK; High-Dimensional Neurology, Institute of Neurology, University College London, London, UK; UCL Medical School, Faculty of Medical Sciences, University College London, London, UK

## Abstract

**Background:** Implantable telemetric intracranial pressure sensors (telesensors) enable routine, non-invasive ICP feedback, aiding clinical decision-making and attribution of pressure-related symptoms in patients with CSF shunt systems. Here, we aim to explore the impact of these devices on service demand and costs in patients with adult hydrocephalus.

**Methods:** We performed an observational propensity matched control study, comparing all patients who had an MScio/Sensor Reservoir (Christoph Mietke, GmbH & Co) against those with a non-telemetric reservoir between March 2016 and March 2018. Patients were matched based on demographics, diagnosis, shunt-type and revision status. Neurosurgical service usage was recorded with frequencies of neurosurgical admissions, outpatient clinics, scans and further surgical procedures in the two years prior and after shunt insertion.

**Results:** 136 patients: 73 telesensor and 63 controls were included in this study (48 matched pairs). Telesensor use led to a significant decrease in neurosurgical inpatient admissions, radiographic encounters and further procedures such as intracranial pressure monitoring. After multivariate adjustment, the mean cumulative saving after two years was £5236 ($6338) in telesensor patients (£5498 on matched pair analysis). On break-even analysis, cost-savings were likely to be achieved within 8 months of clinical use, post-implantation. Telesensor patients also experienced a significant reduction in imaging-associated radiation (4 mSv) over two years.

**Conclusions:** The findings of this exploratory study reveal that telesensor implantation is associated with reduced service demand and provides net financial savings from an institutional perspective. Moreover, telesensor patients required fewer appointments, invasive procedures, and had less radiation exposure, indicating an improvement in both their experience and safety.

- **What is already known on this topic** – Despite shunt insertion being one of the most common treatments for adult hydrocephalus, there is a lack of research concerning the economic viability of shunt components. This includes newer devices such as telemetric sensor reservoirs (telesensors) which can measure intracranial pressure non-invasively, and are posited to have various clinical benefits, but have a greater base cost.
- **What this study adds** – To the best of our knowledge, this is the first matched case-control study evaluating the service demands associated with telesensor use in the largest cohort of this group worldwide. It provides compelling evidence that the integration of telesensors in a diverse set of adult hydrocephalus patients with CSF shunt systems yields noteworthy reductions in service demands, cost and radiation exposure as compared to non-telemetric alternatives.
- **How this study might affect research, practice or policy** – The results of this study will encourage further confirmatory research in financial utility analysis in adult hydrocephalus. It will likely influence practice in supporting a healthcare business model for greater telesensor use.

## Introduction

The implantation of a shunt system represents the principal treatment option for long-term cerebrospinal fluid (CSF) diversion and regulation of intracranial pressure (ICP) in adult hydrocephalus [1]. Incorporating a magnetically adjustable valve in series with the shunt allows for fine-tuning of CSF drainage to regulate ICP [2] and implantable reservoirs allow for percutaneous CSF sampling if necessary. Following shunt insertion, valve adjustments are typically made based on clinical or radiological evidence of CSF under- or over-drainage. However, patient symptom information is often crude and inaccurate [3], and while conventional CT-based imaging can provide evidence of high or low-pressure states, it exposes patients to unnecessary ionising radiation, resource use and additional outpatient appointments.

Telemetric sensors (“telesensors”) are implantable devices that can measure ICP non-invasively. While some telesensors are restricted for short to intermediate periods of use before removal [4], others are permanently implanted. One such latter device is the MScio® (Christoph Miethke GmbH): a telesensor reservoir, which replaces a traditional reservoir and is implanted in series as part of a shunt system. The MScio offers routine ICP measurement and has shown to be useful in both adults and children [5,6], is accurate against the gold standard of bolt-based ICP measurement [7], is MRI compatible [8] and can conveniently be used in clinic and in different patient positions [9].

The advantages of permanent telesensor devices are manifold. Clinically, they provide rapid, non-invasive measurement of ICP, thereby offering an almost instantaneous method to identify and triage patients with abnormal pressure levels. Normal telemetric readings, on the other hand, can reassure both patients and the surgical team that symptoms, if present, are not linked with shunt dysfunction. From a service perspective, the use of permanent telesensor devices like the MScio may reduce neuroimaging, hospital admissions and further clinic appointments, reducing overall service demand. While these latter benefits have been posited, it remains unclear if the MScio is truly cost-effective and whether and when the initial upfront cost can be recouped.

Here, we present our institutional experience regarding the use of MScio telesensors for adult hydrocephalus. We explore the financial impact associated with use of neurosurgical and hospital services, with the goal of determining the breakeven point for achieving net savings. It is our intention to better inform clinicians, trusts, and researchers in their adult hydrocephalus decision-making, through a comprehensive exploratory analysis of our MScio experience at an academic neurosciences centre which, to the best of our knowledge, has the largest volume of telesensor patients worldwide.

## Methods

### Guidelines

Where relevant, this retrospective matched control study was conducted in accordance with Recommendations for Reporting Cost-Effectiveness Analyses by the Panel of Cost-Effectiveness in Health and Medicine [10] .

### Ethics

The local institutional review board approved this study (122-202021-CA), conducted within the context of a service evaluation in the use of telesensor devices at our centre.

### Patients

This study was conducted in a large-volume tertiary neurosciences centre in London, United Kingdom. Included were all patients who were (i) treated for hydrocephalus using a primary or revision CSF diversion technique (ventriculo-peritoneal shunt, lumbo-peritoneal shunt, endoscopic third ventriculostomy with catheter insertion) that involved use of an MScio sensor reservoir; (ii) the telesensor was implanted between March 2016 to 2018 and (iii) could be followed up for a minimum of two years after implantation. Patients who had an MScio reservoir implanted were chosen during a period of clinical equipoise, rather than based on predetermined guidelines or specific to a certain attending.

The two-year window after implantation was selected to allow an adequate period of time to test whether differences existed in use of services and their associated costs. This time period was also selected to prevent overlap with the Covid-19 pandemic period and associated governmental lockdown restrictions which could bias the number and type of clinical encounters. An equal length window before implantation was selected by the study team to ensure a sufficient period, allowing assessment of the control matching process. Control patients were selected based on the aforementioned inclusion criteria but did not have a telemetric reservoir. For the overwhelming majority of controls: a Sprung reservoir (Christoph Miethke GmbH) was implanted.

### Telesensor

The MScio® (Christoph Miethke GmbH, previously ‘Sensor Reservoir’) is a coin-sized device implanted along the tubing or at the angle of the shunt and the tubing at the burr hole site (Figure 1A). A compressible metal membrane is depressed by adjacent CSF when ICP increases. This mechanical stimulation is sensed by a measuring cell and, when in proximity, communicates the real-time pressure measurement telemetrically to a hand-held receiver (Figure 1B).

**Figure 1.**
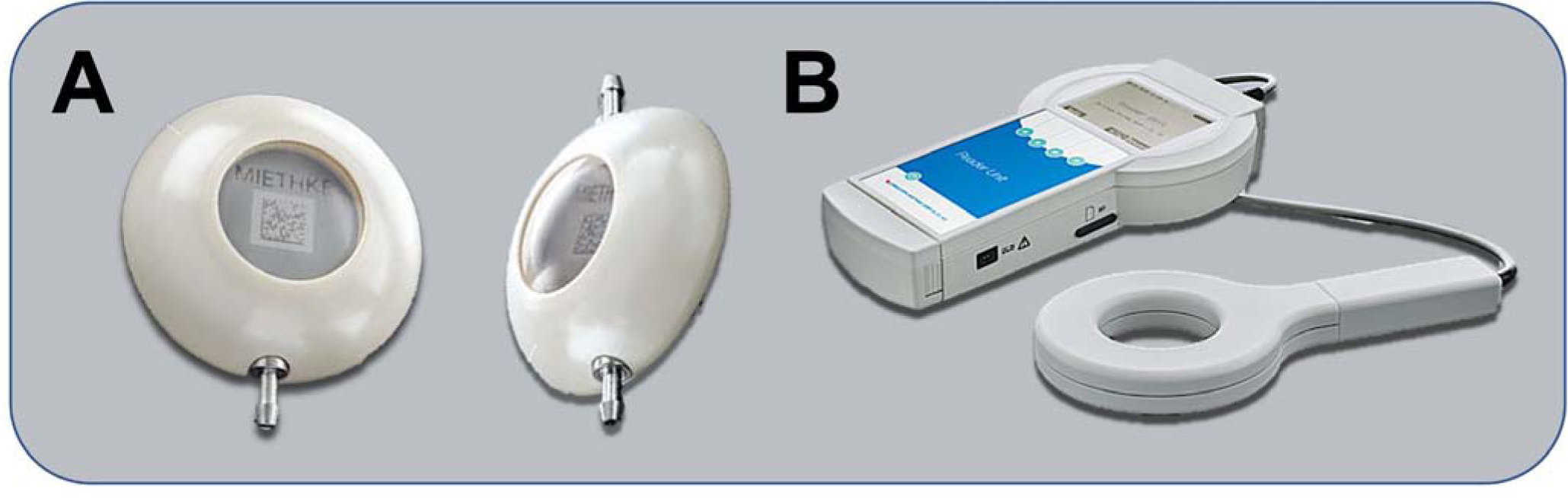
MScio® sensor reservoir and reader system. A: Dome angled MScio implant (left) and in-line MScio implant (right); B: MScio Reader Unit set. Images obtained and adapted with permission from Miethke GmbH.

### Data collection

Data were collected independently by M.A.K., G.R., P.P.S., M.A., Y.A., F.M.Y.L. and P.D.W.B. and double-checked to ensure consistency with A.S.P. Patient demographics, clinical and operative information were retrospectively retrieved from the institution’s electronic health record (Epic System Corporation, Madison Wisconsin, USA) along with the dates and numbers of scans, admissions, and encounters in the period before and after implantation of the telesensor. For neurosurgical encounters, the date, type (admission, outpatient clinic, or correspondence), personnel (attending, resident or nurse specialist), whether the telesensor was read, and any relevant invasive procedures were recorded. For imaging encounters, the date and scan type (MRI, CT, X-ray) were recorded. Encounters relating to the patient’s hydrocephalus condition but were seen by neurology, ophthalmology, and emergency medicine staff were also included. Excluded were same-institution encounters unrelated to hydrocephalus management, shunt insertion or telesensor itself.

### References and tariffs

For calculations in radiation exposure, the following reference values were used: a patient undergoing a plain CT head scan would receive 2.0 mSv, and for an X-ray shunt series of skull, chest and abdomen: 0.9 mSv. For differences in patient costs, calculations were performed based on negotiated local tariffs wherever available for the 2021-22 year. However, these may not be fully reflective nationally or internationally (Table 1). Cost differences were analysed at both annual and bi-annual (total) intervals. USD conversion rates were based on the tariff date of 1/7/22.

**Table 1.**
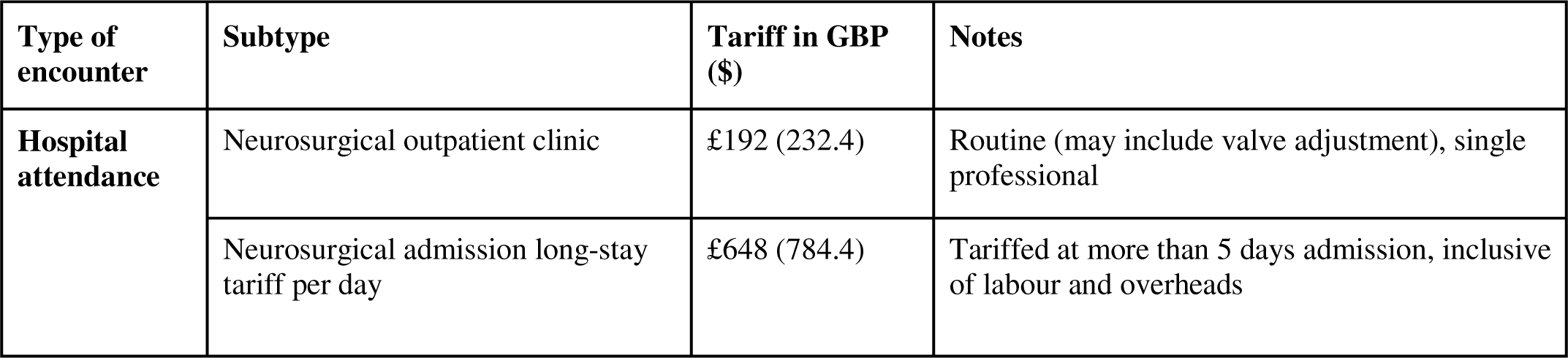
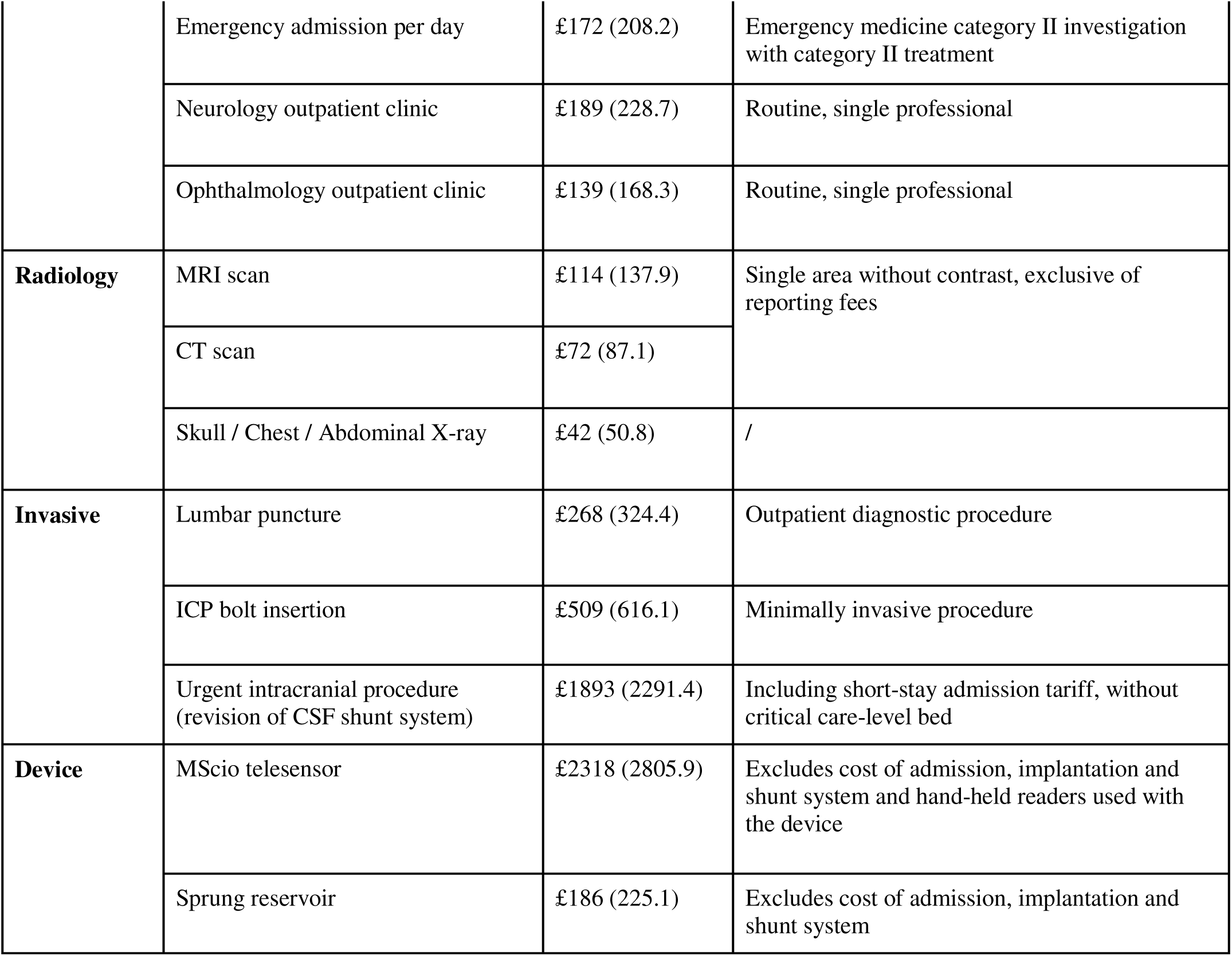
Institutional tariffs for the telesensor device and various hospital encounters inclusive of staff costs. US dollar tariffs based on the foreign exchange rate of £1 = $ 1.2105.

### Matching and data analysis

Matching was performed using a propensity-scoring matching method [11] which aims to pair each telesensor subject with its closest counterpart using a k-nearest neighbours algorithm and propensity logit function based on the following criteria: age, sex, diagnostic category, type of shunt (VPS vs. non-VPS), and whether the patient had previous history of a shunt procedure (see Supplementary Methods for detail). All statistical analyses were performed in Python (v=3.8.1), using pairwise T-tests or Wilcoxon signed rank tests depending on normality testing.

We performed three further analyses. Due to the imperfect nature of the matching process, we performed a sensitivity analysis which would assess two-year financial differences, co-varying the stringency of the matching process to assess if differences would be maintained. Second, because several model variables are likely to interact, we also performed a multivariate linear regression to identify the independent influence of reservoir type on total costs using the complete data set. For simplification, the control group cost was assumed to have a Miethke Sprung reservoir (Table 1). Finally we attempted to find the break-even point at which savings began to be made.

Although the analyses in this paper were exploratory, adjustments (using the Benjamini-Hochberg method) for multiple endpoints are shown where relevant. Our sample size was determined pragmatically based on the total number of telesensor patients available who met the study inclusion criteria in the allocated time period. We performed post-hoc power calculations based on an alpha of 0.05, and effect sizes based on differences in total number of encounters over two years between groups and find that both parametric and non-parametric independent group-wise analyses were sufficiently powered (β > 0.8) [G*Power, v = 3.1].

## Results

### Patient demographics and pre-implant encounter history

136 patients met the inclusion criteria for the study (74 telesensor, 62 controls). Following propensity matching, 48 pairs remained (Table 2). Matched controls were not statistically different from the telesensor group with respect to age, sex, primary diagnosis and type of shunt (Table 2, Supplementary Table 1). When comparing encounters over the 2 years, prior to shunt insertion, control patients were not significantly different with respect to inpatient attendances or invasive procedures (Table 2), however controls did tend to have less neurosurgical outpatient encounters, and significantly less MRI scans (with on average, one less scan per patient in the time period). The full description of the cohort is given in Supplementary Table 2.

**Table 2.**
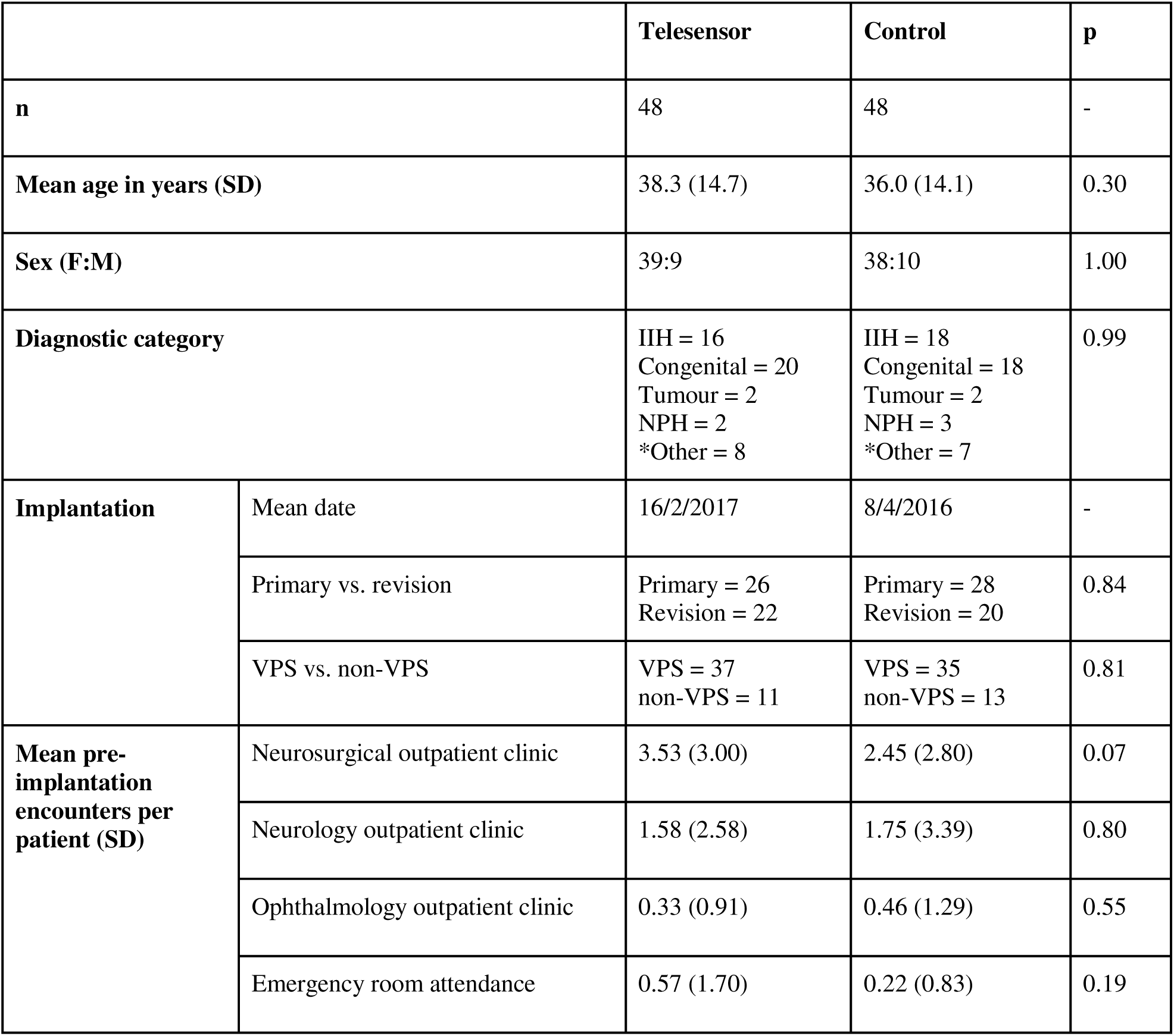
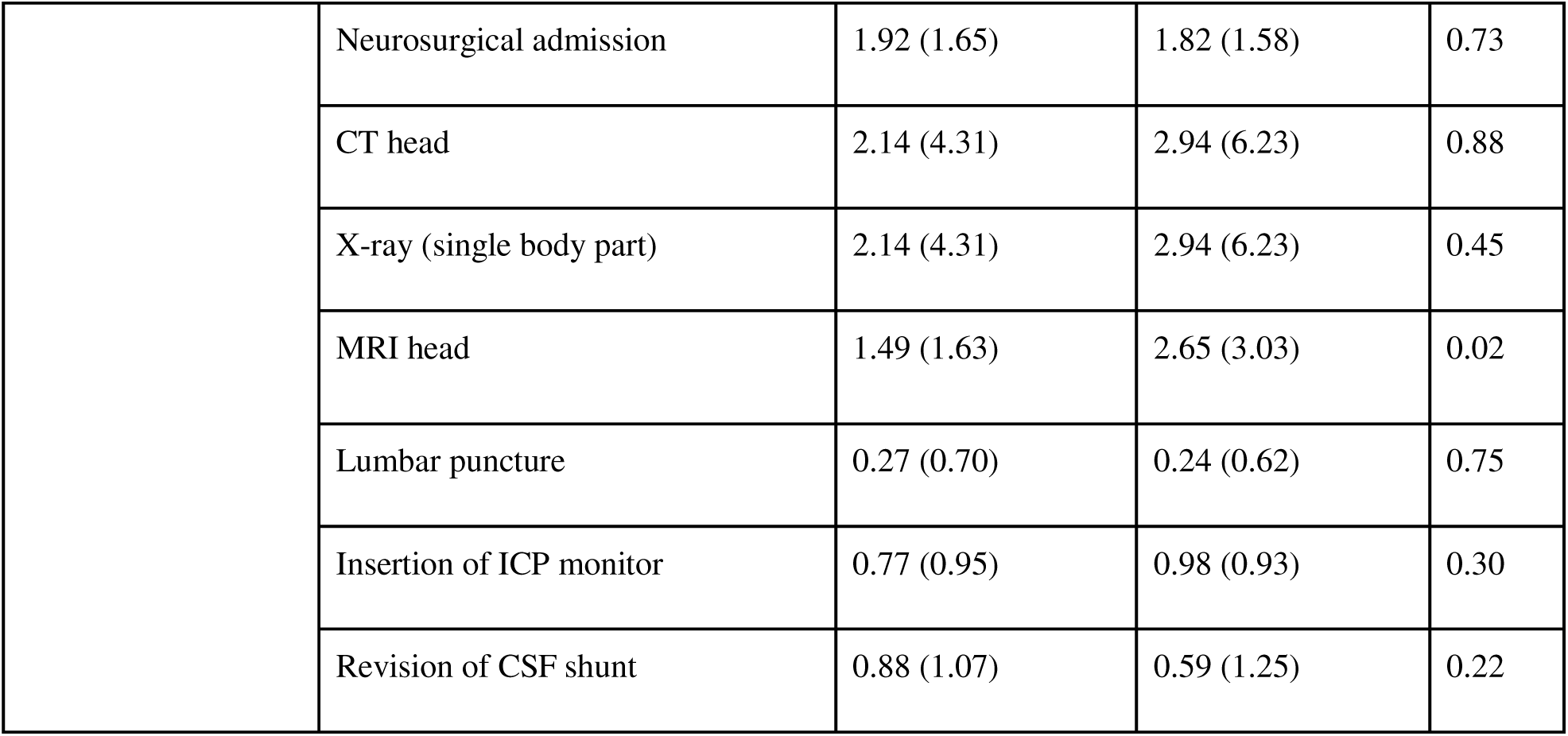
Pre-implantation pairwise comparison between matched telesensor patients and standard reservoir controls with number of encounters evaluated over the preceding two year period. (*Other included CSF leak, secondary hydrocephalus due to subarachnoid haemorrhage, pseudomeningocele and aqueductal stenosis)

### Service demand

The usage of various elective and emergency facilities was evaluated over the 2-year period following implantation (Figure 2, Table 3).

**Figure 2.**
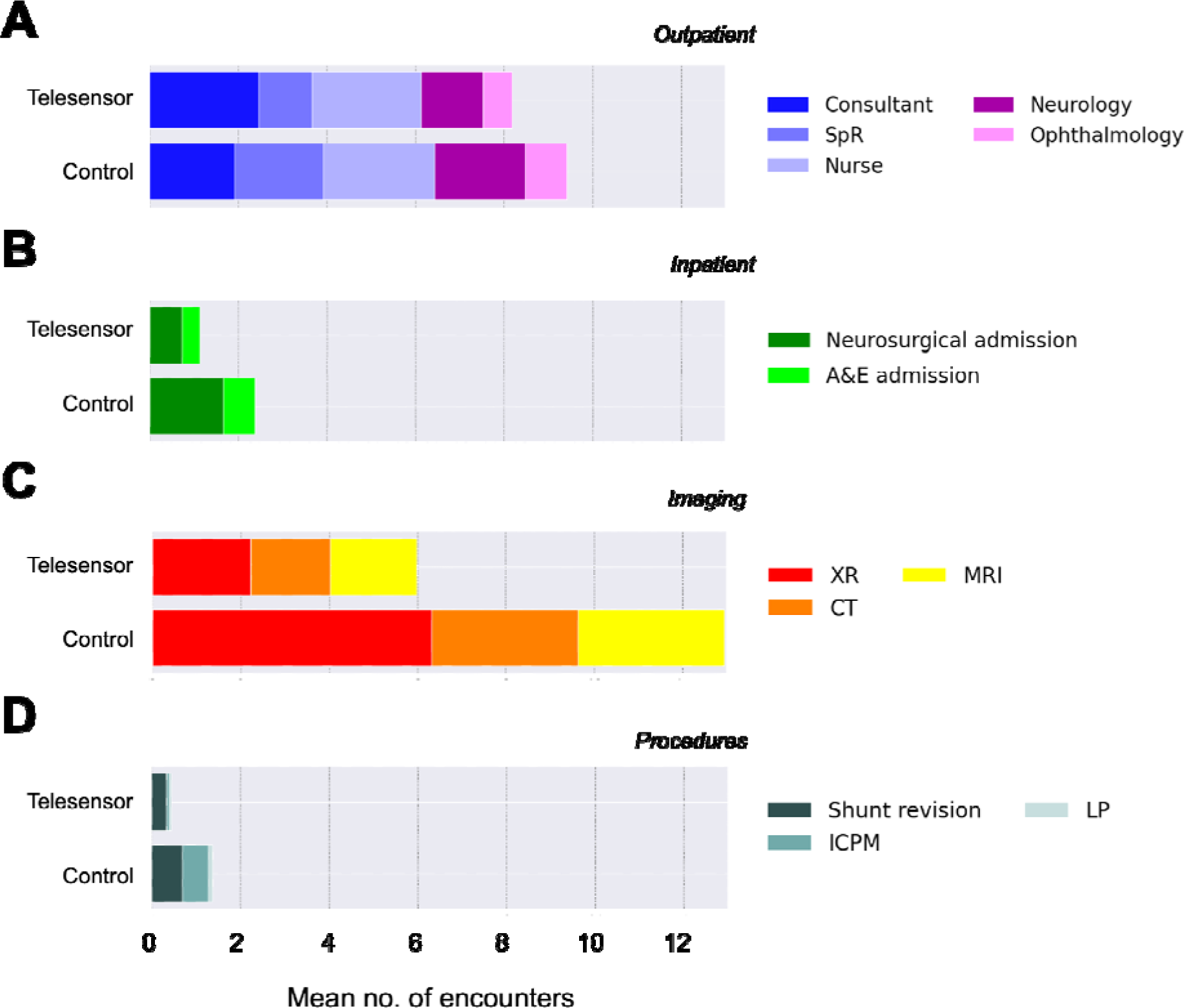
Differences and breakdown of hospital encounters for telesensor and control patients by outpatient attendances (A); inpatient admissions (B); imaging episodes (C); invasive procedures (D).

**Table 3.**
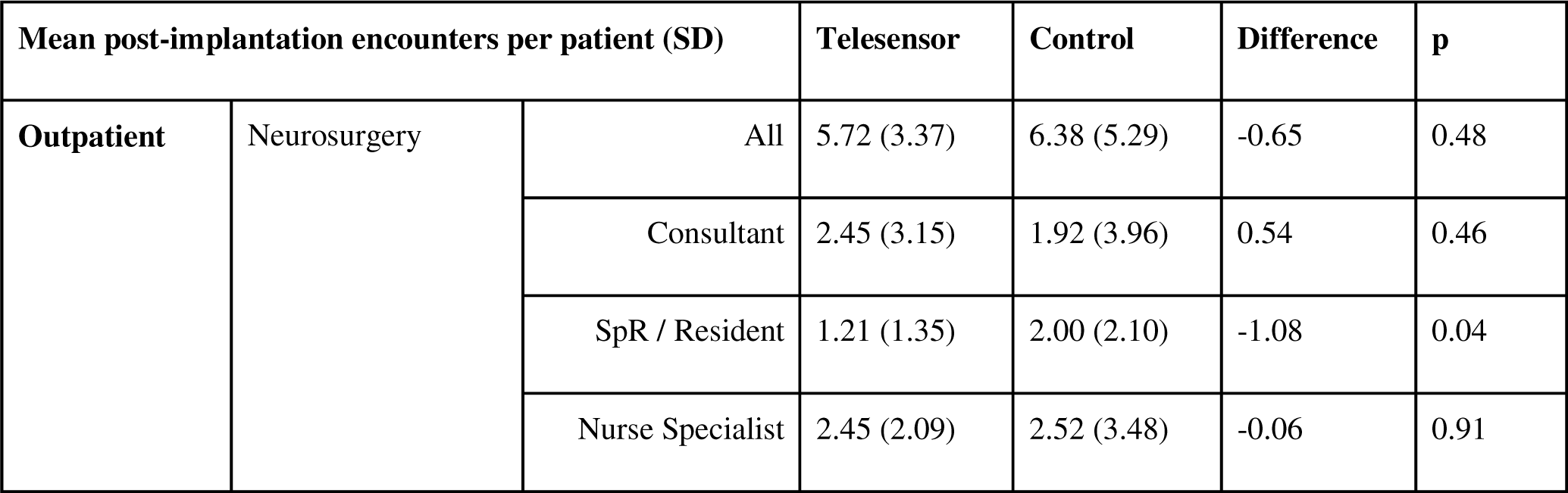
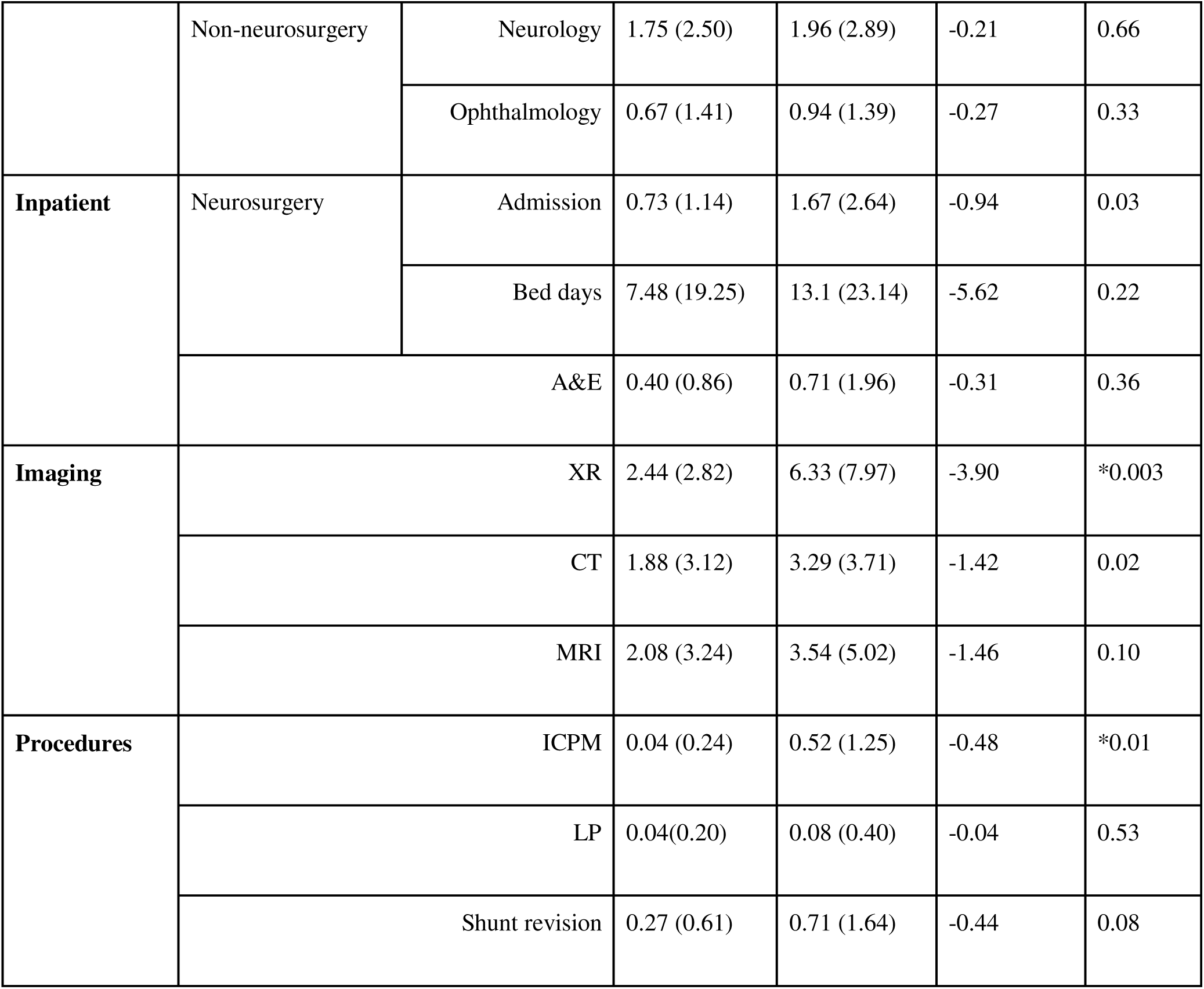
Comparison between patients with telesensors implanted as versus controls who had a standard reservoir with number of pre-implantation encounters evaluated over the preceding two year period. (ICPM = intracranial pressure monitoring; LP = lumbar puncture; SpR = specialist registrar; * would remain significant following multiple comparison adjustment)

#### Outpatient attendance

On average, telesensor patients had 1.97 (SD 2.24) outpatient sensor checks in the two years following implantation and had 1.27 (SD 1.76) outpatient valve adjustments following implantation as compared to controls who had 1.58 (SD 2.07), however this difference was not significant (statistic = w0.77, p = 0.44).

There was no significant difference in the total number of neurosurgery clinic appointments following implantation (between 5 to 6 per patient over 2 years across the cohort); although control patients were more likely to see a registrar (difference = 1.08, statistic = −3.08, p = 0.04). There was no significant difference in neurology or ophthalmology attendances across matched pairs.

#### Inpatient admissions and invasive procedures

Control patients were more likely to require an unprompted neurosurgical admission after the shunt was inserted (difference = 0.94, statistic = 2.23 p = 0.03), and were also more likely to require further intracranial pressure monitoring (difference = 0.48, statistic = −2.57, p = 0.01). Control patients on average had a greater number of inpatient hospital days (13.1 days) as compared to telesensor patients (7.5 days), but this was not found to be significantly different on matched pair analysis. There was also no significant difference between frequency of further lumbar punctures or shunt revisions (although the latter was trending toward more in the control group).

#### Imaging

Telesensor patients had significantly less imaging encounters across all modalities with, on average over two years, 3.9 less single body part X-rays (p = 0.003) and 1.4 less CT head scans (p = 0.02). Based on frequency of imaging encounters -over the two-year period, telesensor patients received approximately, on average, 4.48 mSv (SD 6.64) of radiation, significantly less than control patients who received 8.48 mSv (SD 9.04, p = 0.009), with this difference being equivalent to roughly two years of natural background radiation in the U.K. [12].

### Costs

The difference in costs between telesensor patients and controls during year one, year two and overall are shown in Figure 3 and in Supplementary Table 3. Control patients accrued significantly greater costs related to ICPM, and CT and X-ray imaging in year one and trended towards greater costs for MRI and operative encounters. By the end of year one, the mean pairwise saving (excluding the cost of the implant) was £4624 (p = 0.03). In year two alone, control patients accrued greater costs across almost all the domains however these differences were not significant other than for neurology outpatient appointments (difference = £142, p = 0.04). The mean saving in year two (excluding the cost of the implant) was £874, p = 0.64. When accounting for both years, significant cost differences were found across the same domains as those found in year one. Mean cumulative costs were £7391 among telesensor patients and £12889 among control patients by the end of year two (difference = £5498, p = 0.04). The main burden of cost for both groups was primarily related to neurosurgical inpatient admissions (Figure 3, Supplementary Table 3).

**Figure 3.**
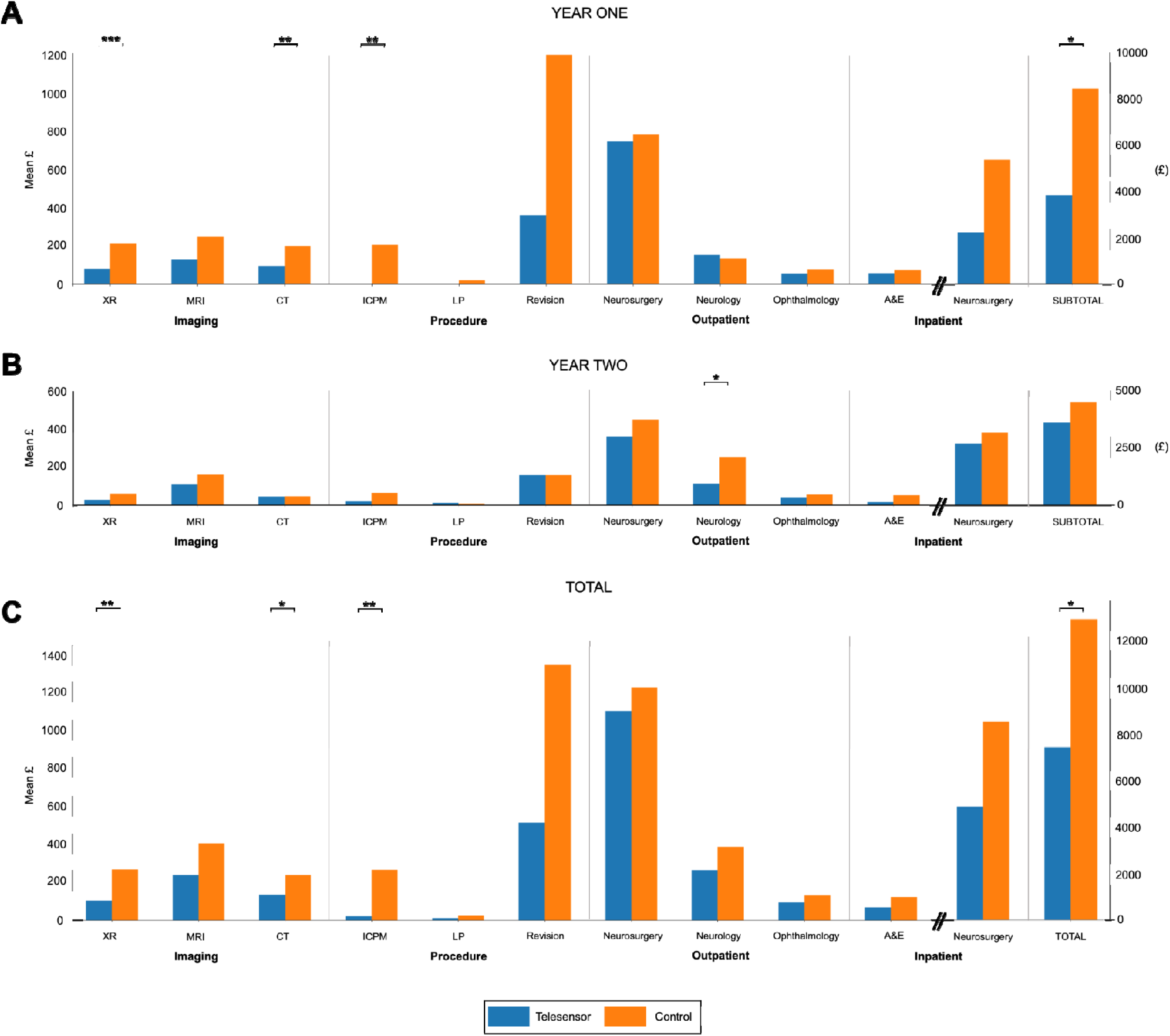
Mean pairwise cost differences between telesensor and control matched pairs at the end of year one (A); year two (B); and overall (C). (* p < 0.05, ** p < 0.01, *** p < 0.001)

### Sensitivity, multivariate and breakeven analyses

We performed a sensitivity analysis to assess whether two-year total costs would differ according to different levels of matching stringency (Supplementary Figure 1). Telesensor savings were maintained across stringency levels with a mean average of £5487 (+/- 1610) across matched pairs, although significant differences were found only up to 50 pairs.

Given that some model variables are likely to interact and patients were excluded during matching, a multivariate regression was performed for the entire data set (Supplementary Table 4). After adjustment, we found that costs associated with telesensor use were significantly less with a saving of £5236 by the end of 2 years (t = −2.05, p = 0.03). Age, sex, and shunt type did not have significant associations in multivariate analyses nor did underlying diagnosis (Supplementary Table 4).

Both pairwise and multivariate analysis did not account for the initial financial burden of the cost of the reservoir. Using the full data set and including for the reservoir base cost, we attempted to identify the point at which a cost saving is made. As a mean average, the break-even point was found to be approximately less than 8 months after shunt insertion (Figure 4)

**Figure 4.**
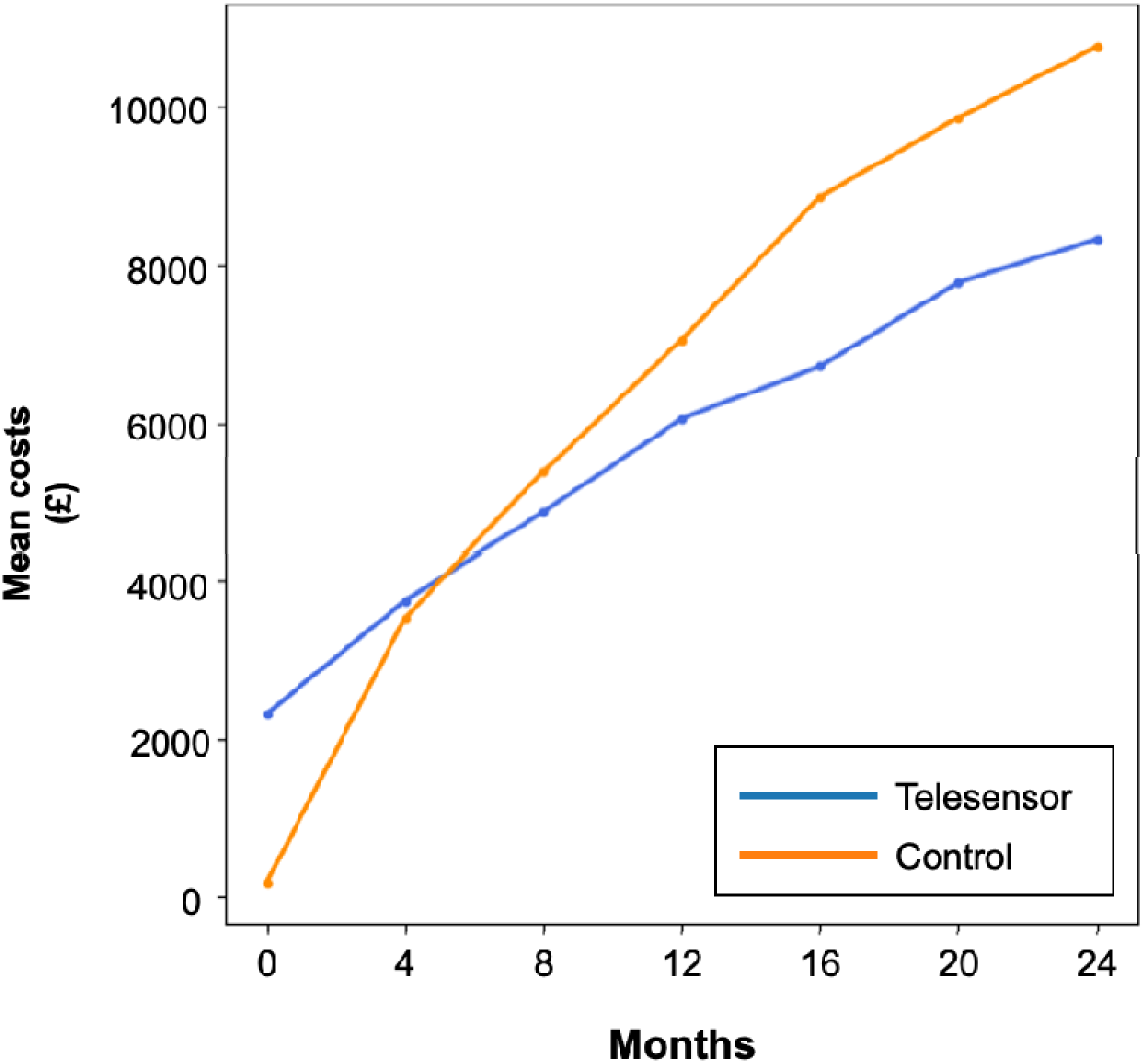
Two-year cumulative mean costs of telesensor and control patients after accounting for initial cost of the shunt reservoir with a break-even point identified at the crossover.

## Discussion

### Summary of results

This observational study assessed service demands and costs in adult hydrocephalus patients requiring CSF shunting who had an implanted telemetric sensor reservoir. Following propensity matching, 48 telesensor patients were compared with 48 controls having non-telemetric reservoirs paired according to demographics, diagnosis and shunt type. Using both univariate or multivariate analyses, on both matched and complete datasets, significant savings were found to be associated with use of telesensor reservoirs. After multivariate adjustment, the mean cumulative saving after two years was found to be £5236 ($6338) when using a telesensor reservoir (£5498 on matched pair analysis). Cost-savings were highly likely to be achieved within 8 months of clinical use after implantation. Finally of specific clinical importance, was the significant reduction in imaging-associated radiation (4 mSv) over two years, for those with a telesensor.

### Interpretation and context

Shunt insertion is a commonly performed adult neurosurgical procedure, with an estimated annual rate of 1,500 patients undergoing primary shunt surgery in the United Kingdom [13]. Research indicates that within the first year, 15% of these patients require revision surgery, predominantly attributable to underdrainage and infection. In spite of the frequency of this surgery, the cost-effectiveness of shunt insertion and the various implantable devices used to assist and control CSF outflow are under-represented within the existing literature.

Kameda et al. conducted a fiscal analysis of shunt surgery for patients with idiopathic normal pressure hydrocephalus (iNPH) [14]. Their work, based on a cohort of 183 Japanese patients using data from SINPHONI and SINPHONI-2 studies, considered various factors such as medical expenses, operation fees, and long-term insurance costs in cost-effective evaluation. The total medical expenses associated with shunt surgery amounted to approximately 12,500$ USD with significant improvements in Quality-Adjusted Life Years (QALYs) compared to non-surgical patients. In a smaller Swedish cohort of 37 iNPH patients, Tullberg et al. used a decision-analytic Markov model to estimate the life-long costs and effects of performing shunt surgery compared to patients receiving no treatment for iNPH [15]. The findings demonstrated that shunt surgery, as a standard treatment for iNPH, resulted in a gain of 1.7 quality-adjusted life years (QALYs) per patient. The cost utility of individual shunt components which are produced by a variety of manufacturers, is less clear. The BASICS trial revealed superior effectiveness of antibiotic-impregnated shunts compared to standard shunts [16]. Here antibiotic shunts were associated with less infections, revisions and were more cost-effective. With respect to shunt valves, in a retrospective single-centre study, Agarwal et al. found that shunt insertion with fixed pressure valves was associated with greater cost savings and similar revision rates to insertion with a programmable valves - although median time of follow-up was around 30 months.

The use of telemetric sensor reservoirs in both adult and paediatric CSF disorders has been increasing over the last decade. While many studies have reported their early experiences and feasibility [17–20] and application in clinical practice [17,21,22], very few have performed a financial-utility analysis - namely, whether the higher base cost of a more sophisticated reservoir translates to better clinical outcomes. In a study conducted by Bjornson et al, a cost-effectiveness analysis of Miethke telesensor reservoirs was performed on a cohort of 12 patients, including 3 children and 9 adults. Here the expenses associated with patient investigations and interventions were analysed during a two-year period prior to, and after, telesensor insertion. Following the telesensor insertion, a significant cost reduction of £6,952 per patient was observed over a two-year duration, along with a corresponding reduction in the frequency of investigations such as CT head scans, X-rays, and ICP monitoring. A larger cost reduction was found as compared to the more modest cost savings found in our work, may at least partially be due to their small heterogeneous sample, which included children, differences in institutional tariffs and that costs were compared with the pre-implantation two years rather than a matched control group.

### Limitations and strengths

We acknowledge a number of limitations in this exploratory study. First, this was a non-randomised retrospective single-centre study with a follow-up period of only two years. Both the matched pair and full cohort had a predisposition toward IIH and congenital hydrocephalus diagnoses. These diagnostic groups, in particular, tend to be high service users as compared to those with acquired hydrocephalus. Although a like-for-like comparison was performed during matching, and diagnosis was regressed as a covariate in multivariate tests; this issue limits the generalisability of the findings to other centres with more heterogeneous patient cohorts and for longer time periods. Second, there was no formal evaluation of ‘effectiveness’ or quality of life. Third, other costs associated with labour and investigations or attendances outside the hospital centre were not collected which may have influenced the results. In spite of these issues, we highlight that our study is the first to formally and comprehensively assess the associated costs and service demands of shunt reservoir use in a large, realistic cohort of both chronic and subacute hydrocephalus patients. We applied a robust statistical design that uses both propensity matching as well as multivariate techniques with results that are remarkably consistent and coherent.

## Conclusion

From an institutional perspective, the implantation of telesensors contributes to a reduction in service demand and a likely net financial saving. From a patient perspective, fewer appointments, invasive procedures, and less radiation exposure suggest an improvement in patient experience and safety. However, further studies are needed to confirm the hypothesis that telesensors are cost-effective in the long-term. Nevertheless, the exploratory findings of this study highlight the potential benefits of telesensors in reducing the financial burden of neurosurgical departments, and their potential to improve the overall management of hydrocephalus.

## Data Availability

All data produced in the present study are available upon reasonable request to the authors, providing ethical permissions have been obtained

## Acknowledgements

The authors personally thank the patients for their time, cooperation, and data used in this publication. We thank all members of the hydrocephalus team at the National Hospital for Neurology and Neurosurgery for their contribution.

## Notes

**Funding:** A.K.T is supported by the NIHR UCLH Biomedical Research Centre, E.M.M. is supported by BBraun. None of the funding sources influenced the design, analysis or reporting of the study.

**Conflicts of interest:** The authors have no conflicts of interest to declare.

### Competing Interest Statement

Funding:
A.K.T is supported by the NIHR UCLH Biomedical Research Centre, E.M.M. is supported by BBraun. None of the funding sources influenced the design, analysis or reporting of the study.
Conflicts of interest:
The authors have no conflicts of interest to declare.

### Funding Statement

This study did not receive any funding

### Author Declarations

The institutional review board (Queen Square Quality and Safety Committee) approved this study (122-202021-CA), conducted within the context of a service evaluation in the use of telesensor devices at our centre.

## References

1 Stagno V, Navarrete EA, Mirone G, et al. Management of Hydrocephalus Around the World. World Neurosurg 2013;79:S23.e17–S23.e20. doi:10.1016/j.wneu.2012.02.004

2 Yamashita N, Kamiya K, Yamada K. Experience with a programmable valve shunt system. J Neurosurg 1999;91:26–31. doi:10.3171/jns.1999.91.1.0026

3 Banks P, Pandit A, Toma A, et al. Happy-ICP: single-centre cohort of 184 telemetric intracranial pressure monitors. Brain Spine 2021;1:100744. doi:10.1016/j.bas.2021.100744

4 Antes S, Tschan CA, Heckelmann M, et al. Telemetric Intracranial Pressure Monitoring with the Raumedic Neurovent P-tel. World Neurosurg 2016;91:133–48. doi:10.1016/j.wneu.2016.03.096

5 Antes S, Stadie A, Müller S, et al. Intracranial Pressure–Guided Shunt Valve Adjustments with the Miethke Sensor Reservoir. World Neurosurg 2018;109:e642–50. doi:10.1016/j.wneu.2017.10.044

6 Freimann FB, Schulz M, Haberl H, et al. Feasibility of telemetric ICP-guided valve adjustments for complex shunt therapy. Child’s Nerv Syst 2014;30:689–97. doi:10.1007/s00381-013-2324-0

7 Adam A, Robison J, Lu J, et al. Abstracts from Hydrocephalus 2016. Fluids Barriers Cns 2017;14:15. doi:10.1186/s12987-017-0054-5

8 Shellock FG, Knebel J, Prat AD. Evaluation of MRI issues for a new neurological implant, the Sensor Reservoir. Magn Reson Imaging 2013;31:1245–50. doi:10.1016/j.mri.2013.03.012

9 Ertl P, Hermann E, Heissler H, et al. Telemetric Intracranial Pressure Recording via a Shunt System Integrated Sensor: A Safety and Feasibility Study. J Neurological Surg Part Central European Neurosugery 2017;78:572–5. doi:10.1055/s-0037-1603632

10 Siegel JE, Weinstein MC, Russell LB, et al. Recommendations for Reporting Cost-effectiveness Analyses. JAMA 1996;276:1339–41. doi:10.1001/jama.1996.03540160061034

11 Kline A, >Luo Y. PsmPy: A Package for Retrospective Cohort Matching in Python. 2022 44th Annu Int Conf IEEE Eng Med Biol Soc (EMBC) 2022;00:1354–7. doi:10.1109/embc48229.2022.9871333

12 Ionising radiation: dose comparisons. https://www.gov.uk/government/publications/ionising-radiation-dose-comparisons/ionising-radiation-dose-comparisons (accessed 9 Jul 2023).

13 Fernández-Méndez R, Richards HK, Seeley HM, et al. Current epidemiology of cerebrospinal fluid shunt surgery in the UK and Ireland (2004–2013). *J Neurol*, Neurosurg Psychiatry 2019;90:747. doi:10.1136/jnnp-2018-319927

14 Kameda M, Yamada S, Atsuchi M, et al. Cost-effectiveness analysis of shunt surgery for idiopathic normal pressure hydrocephalus based on the SINPHONI and SINPHONI-2 trials. Acta Neurochir 2017;159:995–1003. doi:10.1007/s00701-017-3115-2

15 Tullberg M, Persson J, Petersen J, et al. Shunt surgery in idiopathic normal pressure hydrocephalus is cost-effective—a cost utility analysis. Acta Neurochir 2018;160:509–18. doi:10.1007/s00701-017-3394-7

16 Mallucci CL, Jenkinson MD, Conroy EJ, et al. Antibiotic or silver versus standard ventriculoperitoneal shunts (BASICS): a multicentre, single-blinded, randomised trial and economic evaluation. Lancet 2019;394:1530–9. doi:10.1016/s0140-6736(19)31603-4

17 Bjornson A, Henderson D, Lawrence E, et al. The Sensor Reservoir—does it change management? Acta Neurochir 2021;163:1087–95. doi:10.1007/s00701-021-04729-y

18 Norager NH, Lilja-Cyron A, Hansen TS, et al. Deciding on the Appropriate Telemetric Intracranial Pressure Monitoring System. World Neurosurg 2019;126:564–9. doi:10.1016/j.wneu.2019.03.077

19 Müller SJ, Freimann FB, Brelie C von der, et al. Test-Retest Reliability of Outpatient Telemetric Intracranial Pressure Measurements in Shunt-Dependent Patients with Hydrocephalus and Idiopathic Intracranial Hypertension. World Neurosurg 2019;131:e74–80. doi:10.1016/j.wneu.2019.07.014

20 Pennacchietti V, Prinz V, Schaumann A, et al. Single center experiences with telemetric intracranial pressure measurements in patients with CSF circulation disturbances. Acta Neurochir 2020;162:2487–97. doi:10.1007/s00701-020-04421-7

21 Richard KE, Block FR, Weiser RR. First clinical results with a telemetric shunt-integrated ICP-sensor. Neurol Res 1999;21:117–20. doi:10.1080/01616412.1999.11740906

22 Pedersen SH, Norager NH, Lilja-Cyron A, et al. Telemetric intracranial pressure monitoring in children. Child’s Nerv Syst 2020;36:49–58. doi:10.1007/s00381-019-04271-4

