## Supplementary Material for "The impact of intracranial pressure telesensors: an observational propensity matched-control analysis of service demand and costs"

### Supplementary Methods

##### Matching

Matching was performed in a 1:1 format, without replacement, and done iteratively until the distribution of propensity scores between groups were similar and the covariate effect size for all five criteria was less than prior to matching. In this way patients who were unmatched were discarded before formal analysis. Pairwise analysis between telesensor and control patients was then performed in two stages: (i) comparison of service usage and radiation exposure over the two years after implantation and (ii) comparison of annual and total costs.

### Supplementary Results

#### Patient demographics and pre-implant encounter history

**Supplementary Table 1.** Comparison between telesensor patients and controls with respect to specific diagnoses

|  | | Telesensor | Control | p |
| --- | --- | --- | --- | --- |
| Specific diagnosis | | IIH = 16  Congenital = 20  Tumour = 2  Aqueductal stenosis = 3  NPH = 2  Other = 5 | IIH = 18  Congenital = 18  Tumour = 2  Aqueductal stenosis = 3  NPH = 3  Other = 4 | 0.99 |

**Supplementary Table 2.** Demographic characteristics of the full data set

| **n** | | 136 |
| --- | --- | --- |
| **Mean age in years (SD)** | | 39.0 (16.9) |
| **Telesensor:control** | | 74:62 |
| **Sex (F:M)** | | 103:33 |
| **Diagnostic category** | | IIH = 51  Congenital = 43  Tumour = 11  NPH = 14  Other = 17 |
| **Implantation** | Primary vs. revision | Primary = 76  Revision = 60 |
|  | VPS vs. non-VPS | VPS = 105  non-VPS = 31 |

**Supplementary Table 3.** Mean pairwise annual and total cost differences between telesensor and control matched pairs. (* would remain significant following multiple comparison adjustment)

| **Post-implantation encounter, mean cost in GBP (SD)** | | **Year 1** | | | | **Year 2** | | | | **Total** | | | |
| --- | --- | --- | --- | --- | --- | --- | --- | --- | --- | --- | --- | --- | --- |
|  |  | **Telesensor** | **Control** | **Diff** | **p** | **Telesensor** | **Control** | **Diff** | **p** | **Telesensor** | **Control** | **Diff** | **p** |
| **Outpatient** | Neurosurgery | 736 (482) | 772 (701) | -36 | 0.78 | 364 (339) | 452 (414) | -88 | 0..25 | 1100 (648) | 1224 (1017) | -124 | 0.48 |
|  | Neurology | 150 (256) | 130 (203) | 20 | 0.67 | 114 (202) | 256 (437) | -142 | 0.04 | 263 (403) | 386 (542) | -122 | 0.20 |
|  | Ophthalmology | 52 (120) | 75 (134) | -23 | 0.38 | 41 (103) | 55 (98) | -14.5 | 0.42 | 93 (197) | 130 (193) | -38 | 0.33 |
| **Inpatient** | Neurosurgery | 2214 (5967) | 5387 (11968) | -3173 | 0.11 | 2633 (8323) | 3105 (7909) | -472 | 0.78 | 4847 (12475) | 8492 (14998) | -3645 | 0.22 |
|  | A&E | 54 (129) | 72 (209) | -18 | 0.63 | 14 (60) | 50 (154) | -36 | 0.15 | 68 (149) | 122 (338) | -54 | 0.33 |
| **Imaging** | XR | 76 (95) | 207 (245) | -130 | *<0.001 | 26 (63) | 60 (136) | -33 | 0.14 | 102 (118) | 266 (335) | -163 | *0.003 |
|  | CT | 92 (186) | 192 (246) | -101 | *0.009 | 43 (94) | 45 (78) | -2 | 0.93 | 135 (225) | 237 (267) | -102 | 0.03 |
|  | MRI | 126 (170) | 242 (380) | -116 | 0.06 | 112 (247) | 162 (272) | -50 | 0.37 | 238 (369) | 404 (573) | -166 | 0.11 |
| **Procedures** | ICPM | 0 (0) | 201 (489) | -201 | 0.006 | 21 (103) | 64 (200) | -42 | 0.21 | 21 (103) | 265 (639) | -244 | 0.01 |
|  | LP | 0 (0) | 17 (86) | -17 | 0.18 | 11 (54) | 6 (39) | 5 | 0.57 | 11 (54) | 22 (108) | -11 | 0.53 |
|  | Shunt revision | 355 (1007) | 1183 (2912) | -828 | 0.07 | 158 (657) | 158 (657) | 0 | 1.00 | 513 (1154) | 1341 (3098) | -828 | 0.08 |
| **Total** | | 3854 (6096) | 8478 (14718) | -4624 | *0.03 | 3537 (9046) | 4411 (8641) | -874 | 0.64 | 7391 (12963) | 12889 (18516) | -5498 | *0.04 |

**Supplementary Table 4.** Multivariate linear regression with 2-year total cost as the dependent variable

| **Variable** | | **Coefficient (£)** | **SE** | **t** | **p** |
| --- | --- | --- | --- | --- | --- |
| **Intercept** | | 17470 | 4400 | 3.97 | <0.001 |
| **Demographics** | Age | -21 | 88 | -0.24 | 0.81 |
|  | Male | -4272 | 3311 | -1.29 | 0.14 |
|  | Female | reference | | | |
| **Shunt** | Telesensor | -5236 | 2560 | -2.05 | 0.03 |
|  | Control | reference | | | |
|  | Revision | -2677 | 2519 | -1.06 | 0.29 |
| **Diagnosis** | IIH | -3444 | 2968 | -1.16 | 0.25 |
|  | NPH | -2164 | 1538 | -1.41 | 0.13 |
|  | Tumour | -7112 | 4825 | -1.47 | 0.14 |
|  | Other | -3175 | 4046 | -0.785 | 0.43 |
|  | Congenital | reference | | | |

##### **Supplementary Figure 1.** Sensitivity analysis assessing influence of matching stringency on two-year costs for telesensor and control matched pairs. (* p<0.05; ** p<0.01)

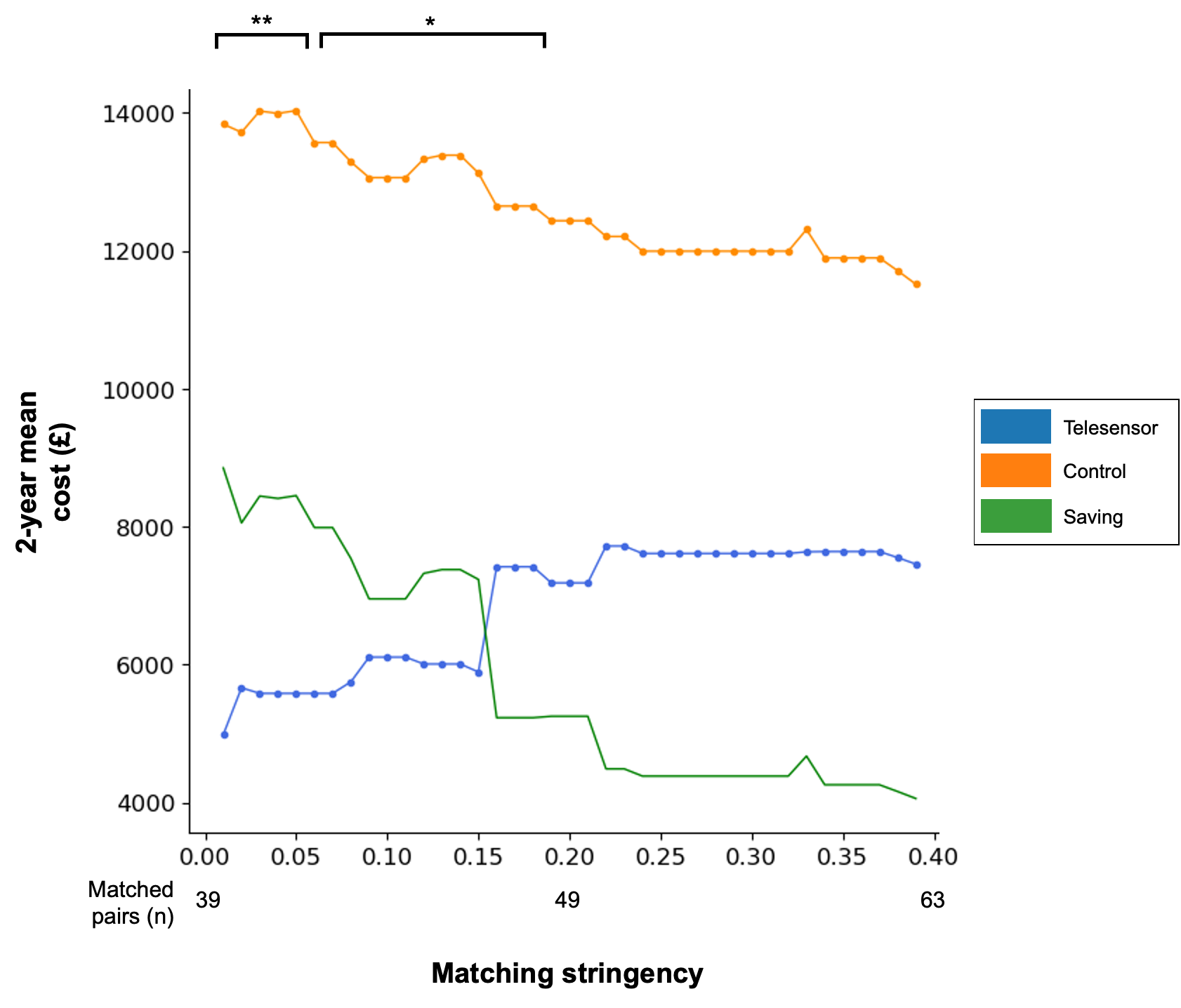
